# A Patient-Specific Framework for Separating Temporal Localization from Intracranial Waveform Reconstruction of Interictal Epileptiform Discharges from Scalp EEG

**DOI:** 10.64898/2026.09.25.26364055

**Authors:** Teppei Matsubara, Ryu Koda, Mark Richardson, Steven Stufflebeam

**Affiliations:** Athinoula A. Martinos Center for Biomedical Imaging, Department of Radiology, Massachusetts General Hospital, Boston, MA, USA; Harvard Medical School, Boston, MA, USA; Tohoku University School of Medicine, Sendai, Japan; Department of Neurosurgery, Massachusetts General Hospital, Boston, MA, USA

**Keywords:** intracranial electroencephalography, scalp electroencephalography, interictal epileptiform discharge, patient-specific models, deep learning

## Abstract

**Objective:** We developed a patient-specific framework to determine which information about interictal epileptiform discharges (IEDs) can be recovered from scalp EEG, separating temporal localization (“where”) from event-specific intracranial waveform reconstruction (“what”).

**Methods:** Six subjects from a public simultaneous high-density scalp EEG and intracranial EEG (iEEG) dataset were analyzed using patient-specific dual-branch convolutional-attention models integrating waveform/topographic and time-frequency representations. For the where task, a 640-ms scalp EEG segment was divided into eight 80-ms bins, and the model predicted the bin containing the intracranial IED peak. For the what task, a 250-ms peak-centered iEEG waveform was represented as a fixed patient-specific template plus an event-specific residual predicted from scalp EEG. Direct scalp-input controls and event-clustered bootstrap confidence intervals were used to distinguish scalp-dependent information from template-driven performance.

**Results:** Where accuracy varied markedly across subjects (0.080–0.984), with an unweighted subject-level mean of 0.333 versus a chance level of 0.125; 95% confidence intervals were above chance level in three of six subjects. Input-control analyses supported a scalp-dependent contribution in subjects with successful localization. For the what task, mean waveform correlation was 0.752 for the model and 0.754 for the fixed patient-specific template (mean difference,-0.002), while residual correlations were generally modest (−0.020 to 0.378).

**Conclusion:** Scalp EEG contained recoverable temporal information in some subjects, but event-specific single-contact morphology was not consistently recovered beyond the patient-specific template. Separating temporal localization from template-based waveform reconstruction provides a more stringent framework for evaluating noninvasive-to-intracranial inference and reduces conflation of prior-driven similarity with input-dependent reconstruction.

## 1. Introduction

Interictal epileptiform discharges (IEDs) are central electrophysiological biomarkers in the presurgical evaluation of drug-resistant epilepsy (Kural et al., 2020). Intracranial electroencephalography (iEEG), including stereo-EEG, provides high-spatiotemporal-resolution invasive recordings of IEDs and is often required when noninvasive studies are insufficient to define the epileptogenic zone (Rosenow and Luders, 2001). In contrast, scalp EEG is noninvasive, widely available, and routinely used in epilepsy evaluation, but many intracranial IEDs are not clearly visible at the single-trial scalp level because of attenuation by intervening tissues, source depth, spatial cancellation, and limited scalp signal-to-noise ratio (SNR). Scalp visibility also depends on the spatial extent and synchrony of the underlying cortical discharge (Tao et al., 2005). Simultaneous scalp EEG and iEEG recordings provide a unique opportunity to investigate what information about intracranial IEDs remains observable noninvasively (Ramantani et al., 2016). Previous simultaneous recordings have shown that intracranially detected IEDs that are difficult to identify in individual scalp EEG events can nevertheless produce reproducible scalp responses after event-locked averaging (Zauli et al., 2024). Computational analyses have further demonstrated that scalp signatures of intracranial IEDs may be recoverable even when the events are not visually apparent (Pyrzowski et al., 2021; Spyrou et al., 2016).

Most machine-learning studies of IEDs have focused on detection—determining whether and when an IED occurs in scalp EEG. Recent deep-learning approaches, including SpikeNet2 (Li et al., 2025) and patient-specific detection frameworks (Jing et al., 2020), have demonstrated increasingly accurate identification of IEDs. More recently, machine-learning analysis of simultaneous scalp and intracranial EEG has identified scalp markers associated with mesial temporal IEDs that were not visually detectable on scalp EEG (Yamaguchi et al., 2026). Detection is essential for quantifying discharge occurrence and is a prerequisite for practical analysis of continuous EEG. However, detection alone does not address whether scalp EEG also contains information about the intracranial morphology associated with an individual event. This question may be relevant for longitudinal patient-specific monitoring, because changes in IEDs may involve not only discharge frequency but also changes in waveform morphology, for example between a spike (20–80 ms) and a broader sharp wave (80–200 ms) (Janmohamed et al., 2022).

A complementary line of work has therefore investigated EEG-to-iEEG signal reconstruction. In particular, E2SGAN demonstrated that generative models can synthesize iEEG-like signals from simultaneously recorded scalp EEG using time-frequency representations and adversarial learning (Hu et al., 2022). This work established the feasibility of noninvasive-to-invasive signal translation, but primarily addressed continuous or segment-level signal synthesis. Whether scalp EEG contains sufficient information to recover the morphology of a single intracranial IED event, particularly when IED morphology is strongly patient-specific, remains less clear. A further difficulty is that within a patient, characteristic intracranial IEDs can be highly stereotyped. Consequently, a model may appear to reconstruct an individual event simply by reproducing a patient-specific average waveform, even if little information is extracted from the scalp EEG itself.

We therefore formulated the problem into two complementary questions: where and what. Given a local scalp EEG segment containing the annotated intracranial event, we first asked whether scalp EEG could identify where in time the primary intracranial IED occurred within that segment. We then asked what intracranial waveform morphology could be reconstructed at the identified event. Using a public simultaneous high-density scalp EEG-iEEG dataset, we developed patient-specific models for these two tasks and performed direct scalp-input ablations to distinguish scalp-dependent information from template-driven reconstruction. Because the available dataset contains only six subjects, one representative iEEG contact and one characteristic IED type per subject, the present study evaluates a patient-specific methodological framework for determining which aspects of intracranial IED information are recoverable from scalp EEG, rather than a continuous IED detector or a cross-patient intracranial signal reconstruction system.

## 2. Materials and Methods

### 2.1. Dataset and Study Design

We used a publicly available simultaneous high-density scalp EEG and stereo-EEG recordings from patients with epilepsy (https://osf.io/89ndr/) (Zauli et al., 2024). Recordings were sampled at 1 kHz and high-pass filtered at 0.5 Hz. The released dataset contains 256-channel scalp EEG together with a representative iEEG contact per subject, selected by Zauli et al. to represent the characteristic IED type. We analyzed six subjects (sub-01, sub-02, sub-04, sub-05, sub-06 and sub-08). Subjects sub-03 and 07 were excluded because multiple epochs contained saturated IED waveforms.

Each event was provided as a 2-s epoch centered near a manually annotated intracranial IED peak. Because the timing of the provided manual annotations showed small event-to-event offsets from the principal iEEG peak, we visually reviewed each event and manually adjusted the peak time when necessary. Hereafter, “IED peak” refers to this corrected peak time. Although additional non-centered spike-like deflections were present in some epochs, the released data do not provide sufficient continuous clinical context to determine reliably whether these additional deflections represent the same IED type, another epileptiform pattern, or non-epileptiform activity. We retained the provided identification of the primary IED but did not relabel additional within-epoch deflections. Accordingly, the present study does not evaluate IED detection in continuous EEG; rather, it evaluates information recoverable from a local scalp EEG segment already known to contain the annotated intracranial event.

The dataset imposes two additional constraints on the present analysis. First, each subject contributes one characteristic IED type, resulting in relatively stereotyped intracranial morphology within subjects and therefore a strong patient-specific waveform prior. Second, only one representative intracranial contact is used as a reconstruction target, whereas scalp EEG contains multichannel spatial information. Thus, similarity of the waveform recorded at a single intracranial contact does not establish that the spatial distribution of the underlying intracranial generator is identical across events. We therefore designed the analysis to separate two questions: where, whether scalp EEG contains information about the temporal position of the primary intracranial IED within a local segment, and what, whether scalp EEG contains information about its event-specific intracranial waveform beyond a fixed patient-specific template.

### 2.2. Temporal Localization and Waveform Reconstruction Problem Formulation

For the where task, scalp EEG was represented as a 640-ms local segment divided into eight non-overlapping 80-ms bins. An 80-ms bin width was chosen as a clinically interpretable temporal resolution corresponding to the upper duration of an epileptiform spike (20–80 ms), whereas sharp waves are typically longer (80–200 ms) (Janmohamed et al., 2022). The target was the bin containing the iEEG peak, formulated as an eight-class classification problem. Local segments were generated at different temporal offsets relative to the IED peak, allowing the peak to occupy different positions among the eight bins rather than being fixed at the center. Thus, the model was required to infer the temporal position of the intracranial event from scalp EEG within a segment already known to contain the event. This task does not constitute detection from arbitrary or continuous EEG.

For the what task, the target was a 250-ms iEEG waveform centered on the IED peak. Because intracranial IED morphology can be strongly patient-specific, we explicitly represented the target as a fixed patient-specific template plus an event-specific residual. The template was constructed using training events only, and the model predicted the residual relative to the fixed template. This formulation allows us to quantify separately the performance attributable to the patient-specific waveform prior and the additional event-specific information contributed by scalp EEG.

All analyses were patient-specific. Events were separated into training, validation, and held-out test sets, and no test event was used to construct the patient-specific template or train the model. Performance therefore represents generalization to new events from the same patient, not generalization across patients.

### 2.3. Patient-Specific Model

A separate model was trained for each subject. For each of the eight 80-ms bins, multichannel scalp EEG was represented by two complementary branches. The waveform branch projected the scalp signals onto a 32×32 topographic grid, preserving both temporal waveform information and scalp spatial distribution. The time-frequency branch represented scalp activity using band-limited topographic maps. The two inputs were separately encoded by convolutional neural networks and concatenated to form a bin-level representation. The fused representation was projected to a common embedding space, and a learnable positional embedding was added to preserve the temporal order of the eight bins. The resulting sequence was processed by a Transformer encoder with an embedding dimension of 128, four attention heads, one layer, and dropout of 0.1. Self-attention allows each temporal-bin representation to incorporate information from the other bins in the sequence, enabling the model to capture dependencies across the 640-ms local temporal context. (Vaswani et al., 2017).

The model was trained in two stages. In the first, where stage, the waveform and time-frequency encoders, temporal-context module, and localization head were optimized to predict which of eight bins contained the IED peak. The localization head produced one logit per bin, which was converted to a probability distribution using a softmax function. To reduce sensitivity to placement of the IED peak near a bin boundary, the training target was represented as a Gaussian-weighted soft label centered on the true peak-containing bin, with a standard deviation of 0.8 bin and support extending to the immediately adjacent bins. The where-stage loss was soft-label cross-entropy. Exact-bin accuracy was used for the final test evaluation.

In the second, what stage, the pretrained waveform and time-frequency encoders, temporal-context module, and where head were frozen, and the residual-waveform head was trained separately. During this stage, contextual representations across the eight bins were pooled using a teacher-forced soft distribution centered on the true peak-containing bin. The reconstruction target was the 250-ms iEEG waveform centered on the IED peak. This waveform was represented as a fixed patient-specific template, derived exclusively from training iEEG events, plus an event-specific residual predicted from scalp EEG. The reconstructed waveform was therefore defined as the sum of the fixed template and the predicted residual.

The what-stage objective was designed to prioritize event-specific deviations from the patient-specific template. It consisted of the mean-squared error between predicted and true residual waveforms, an additional mean-squared-error term emphasizing the central portion of the residual waveform surrounding the IED peak, and a residual-shrinkage penalty discouraging unnecessarily large deviations from the fixed template. The respective loss weights were 0.5, 0.1, and 0.05. No additional full reconstructed-waveform loss was applied. During end-to-end test evaluation, teacher forcing was not used. Instead, contextual representations were pooled according to the probability distribution predicted by the where stage. Thus, uncertainty or error in temporal localization was propagated directly to the primary waveform-reconstruction result. The teacher-forced modes were retained only as auxiliary analyses of reconstruction performance when temporal localization was supplied.

The where and what stages were trained for up to 30 epochs each using the Adam optimizer, with a learning rate of 10^-4^ and weight decay of 10^-5^, and a batch size of 8. Early stopping was applied separately to the two stages with a patience of eight epochs, and the model state with the best validation performance was retained. The held-out test set was not used for training or model selection. The training procedure explicitly optimized the where stage first, restored its best validation state, and subsequently trained the what stage with the where/context components frozen.

### 2.4. Evaluation, Ablation, and Statistical Analysis

The two tasks were evaluated separately on held-out test events. For the where task, performance was quantified as the classification accuracy for identifying the 80-ms bin containing the intracranial IED peak. Each test event was evaluated with the annotated peak placed at each of the eight possible positions within the local segment, yielding a balanced target distribution across bins. Accordingly, both the random-chance and empirical-prior baselines were 1/8 = 0.125.

To determine whether temporal localization depended on scalp EEG rather than positional or dataset structure, we performed direct input-control analyses. These included zeroing both scalp input branches, separately zeroing the waveform or time-frequency branch, shuffling the correspondence between scalp input and events, and disrupting temporal order by shuffling the eight input bins. Performance was also compared with fixed-position baselines. These controls were used to assess whether successful localization required information contained in the scalp EEG input.

For the what task, Pearson correlation between the reconstructed and true iEEG waveform was the primary metric. The same target waveform was also compared with the fixed patient-specific training template. We therefore report the difference in waveform correlation between the model and fixed template, together with the correlation between the predicted and true residual waveforms. This evaluation directly quantifies whether scalp-conditioned residual prediction improves reconstruction beyond the fixed patient-specific waveform prior. Because each source event generated multiple test chunks, observations derived from the same event were not treated as statistically independent. For each subject and analysis condition, 95% confidence intervals were estimated using 2,000 event-clustered bootstrap resamples. Source events were sampled with replacement, and all chunks derived from each sampled event were retained together within a bootstrap sample. Confidence limits were defined by the 2.5^th^ and 97.5^th^ percentiles of the bootstrap distribution. Subject-level summaries across the six patients were calculated without weighting by the number of events. Finally, we explored whether intersubject variability was related to signal characteristics. For each subject, scalp EEG epochs were aligned to the IED peak and averaged across events. For descriptive visualization and SNR analysis, a representative scalp channel was selected as the channel showing the largest peak-to-peak amplitude in the IED-peak-aligned, all-event average within ±250 ms of the IED peak. This channel selection was used only for post hoc descriptive analysis and was not used for model training or model selection. Scalp EEG evoked SNR was then calculated as the peak-to-peak amplitude of the event-averaged response within ±150 ms of the IED peak divided by the root mean square of the event-averaged baseline measured 0.5–0.9 s before and after the peak. iEEG waveform stereotypy was quantified using a leave-one-out procedure. For each annotated IED, a reference waveform was constructed by averaging all other IED events from the same subject, and Pearson correlation between the held-out event and this reference waveform was calculated within the same 250-ms peak-centered window for the what target. Subject-level stereotypy was defined as the median of these event-wise correlations. The fraction of events with negative correlation was additionally calculated as the proportion of opposite-polarity events. Because only six subjects were available, these analyses were considered exploratory and were not used for formal statistical inference.

## 3. Results

### 3.1. Dataset Characteristics and Representative Events

The number of available events varied across subjects, ranging from 36 to 151, with 6–22 held-out test events per subject (Table 1). Event-locked averaging revealed substantial intersubject differences in the visibility of the intracranial IED on scalp EEG (Fig. 1). In contrast, the intracranial waveform at the representative contact was generally stereotyped within each subject, although appreciable event-to-event variability remained.

**Figure 1.**
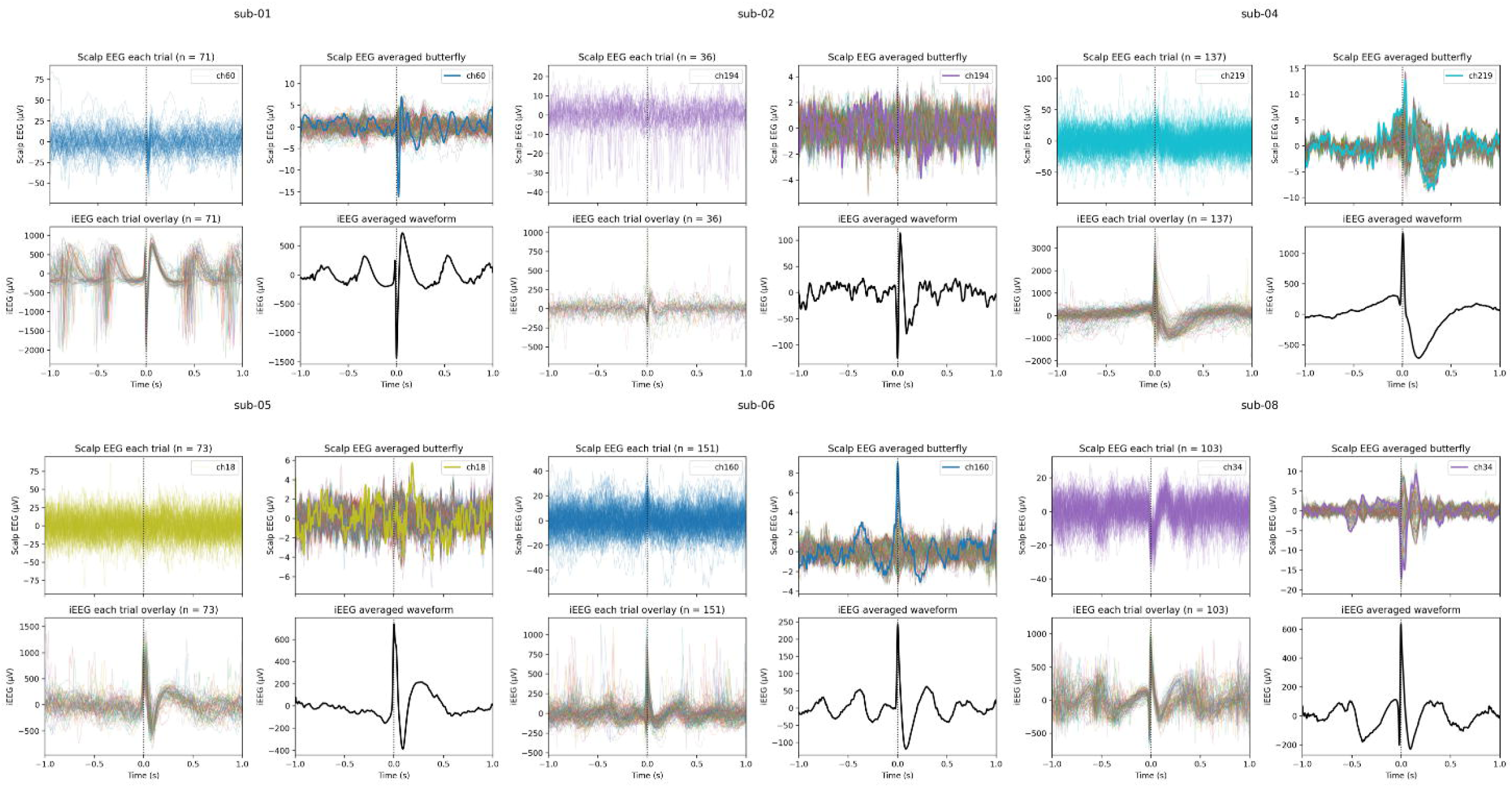
Dataset characteristics across the six subjects. Signals are aligned to the interictal epileptiform discharge (IED) peak (time 0). For each subject, the left panels show individual-event overlays and the right panels show event-averaged responses. The upper panels show scalp EEG, with a representative scalp channel highlighted, and the lower panels show iEEG from one contact. The figure demonstrates substantial intersubject variability in scalp visibility and generally stereotyped, but not identical, intracranial IED morphology. The public dataset is limited to event-centered 2-s epochs, contains additional non-annotated spike-like deflections in some epochs, represents one characteristic IED type per subject, and provides only one representative iEEG contact per subject.

**Table 1.**
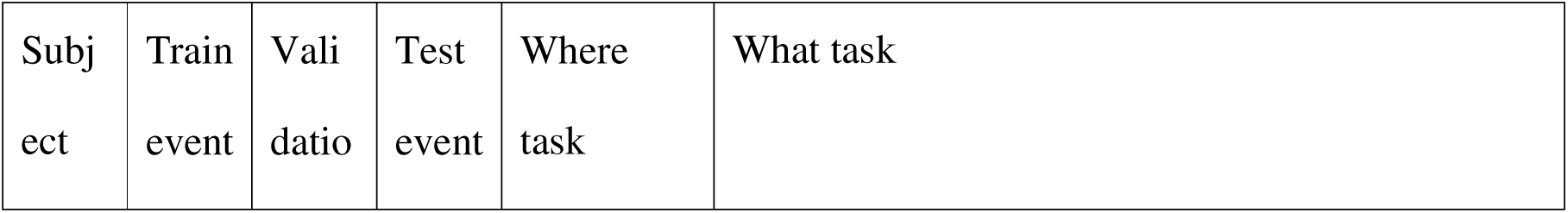
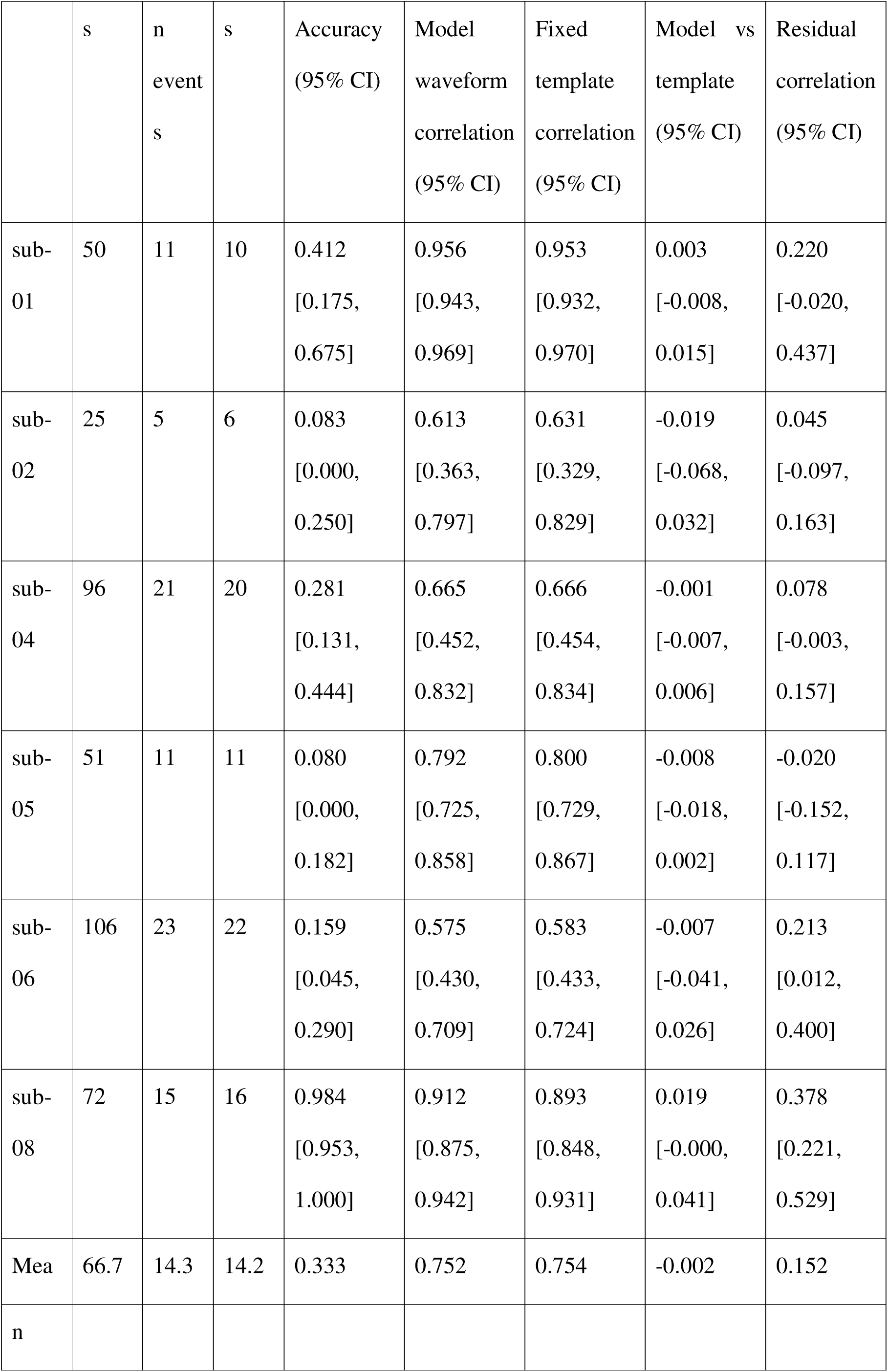
Where and what task performance.

Figure 2 shows illustrative held-out events from all six subjects, including the event with the largest model-template correlation difference and the event with the highest model waveform correlation for each subject. In many events, the model reconstruction closely followed the fixed patient-specific template, whereas event-specific deviations from the template were not consistently reproduced. The examples also illustrate substantial heterogeneity in reconstruction behavior; for example, one sub-04 event showed an opposite-polarity intracranial deflection that was not reproduced by either the model or the fixed template. These observations motivated separate quantitative evaluation of temporal localization and waveform reconstruction.

**Figure 2.**
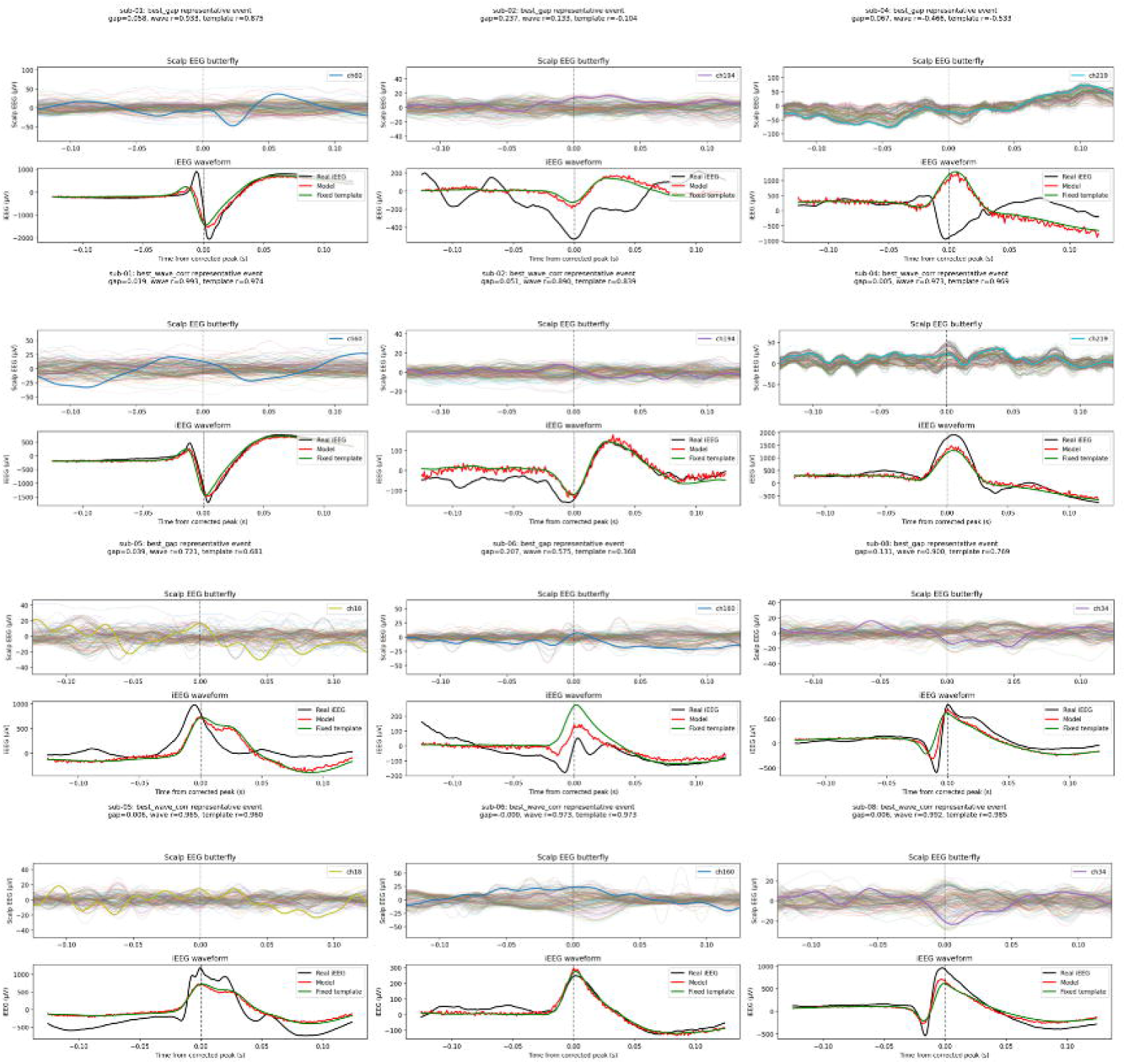
Illustrative single-event intracranial waveform reconstruction across all six subjects. For each subject, two held-out test events are shown: the event with the largest model-template waveform-correlation difference and the event with the highest model waveform correlation. For each event, the upper panel shows the scalp EEG around the IED peak, with a representative channel highlighted, and the lower panels show the real iEEG waveform, model reconstruction, and fixed patient-specific template. Event-specific deviations from the template are not consistently reproduced. The sub-04 example also illustrates an opposite-polarity intracranial event that was not recovered by either the model or the fixed template.

3.2. Temporal Localization: “Where”

Where accuracy varied markedly across subjects (Fig. 3A, Table 1). Accuracy was 0.412 in sub-01, 0.083 in sub-02, 0.281 in sub-04, 0.080 in sub-05, 0.159 in sub-06, and 0.984 in sub-08, compared with a chance level of 0.125. The 95% confidence intervals were entirely above chance for sub-01, sub-04, and sub-08, whereas performance in the remaining subjects was near or below chance. Thus, temporal information about the intracranial IED was recoverable from scalp EEG in some, but not all, subjects.

**Figure 3.**
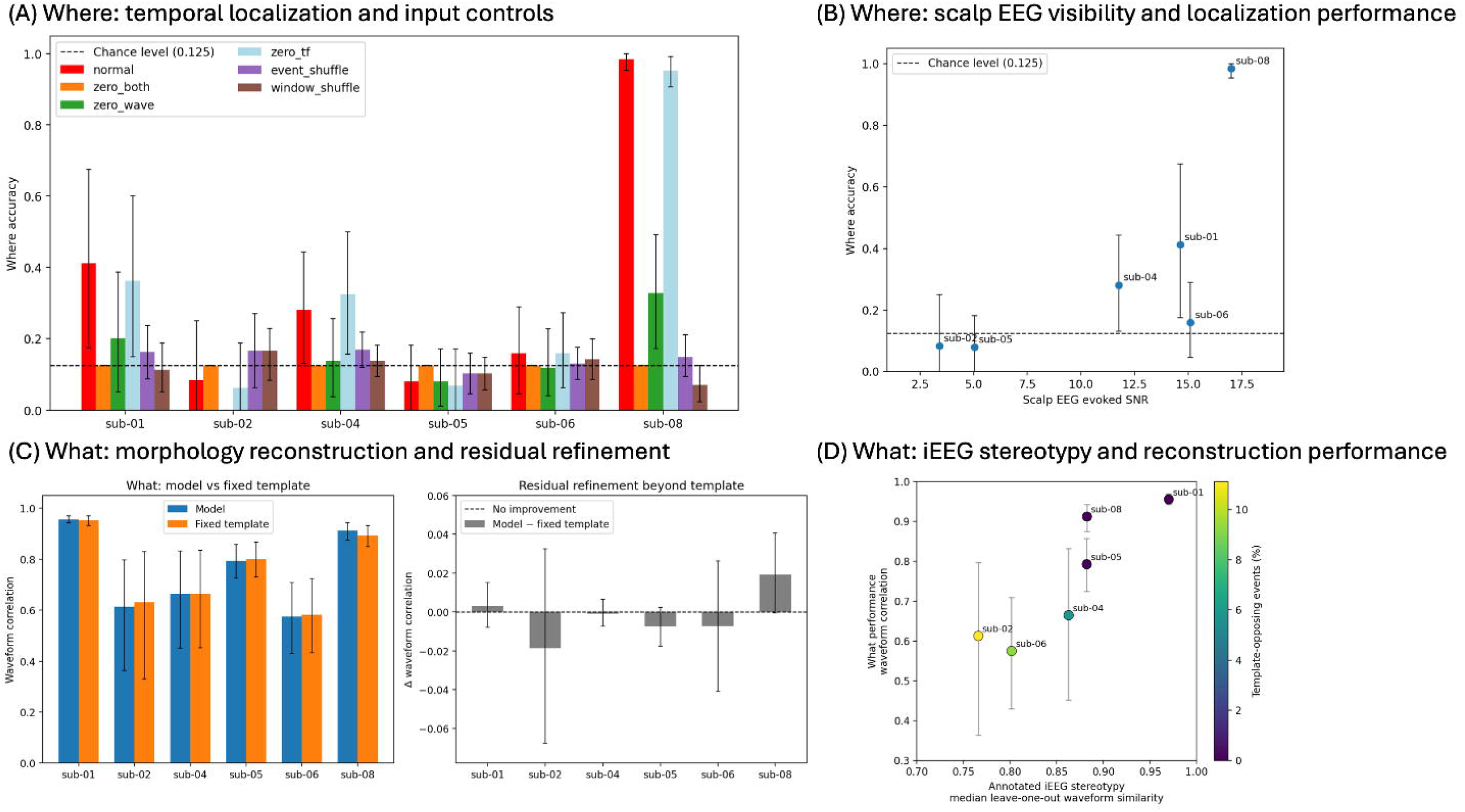
Subject-level performance and factors associated with the where and what tasks. (A) Where: temporal localization accuracy and input controls. The dashed line indicates chance accuracy (1/8 = 0.125). Controls include zeroing both input branches, separately zeroing the waveform or time-frequency branch, event shuffling, and temporal-bin shuffling. Error bars indicate 95% event-clustered bootstrap confidence intervals. (B) Where: relationship between event-locked scalp EEG SNR and localization accuracy. (C) What: model and fixed-template waveform correlations and residual refinement beyond the fixed template. (D) What: relationship between iEEG waveform stereotypy and reconstruction performance. Point color indicates the proportion of template-opposing events, defined as annotated events with negative leave-one-out waveform correlation.

Input-control analyses supported a scalp-dependent contribution in subjects with successful localization (Fig. 3A). In sub-01 and sub-08, zeroing both input branches reduced accuracy to chance, and event shuffling or temporal-bin shuffling markedly degraded performance. Similar degradation was observed in sub-04. Removing the waveform branch generally impaired localization more strongly than removing the time-frequency branch in these subjects, suggesting that temporal localization was driven primarily by information contained in the scalp waveform representation.

Where accuracy also tended to increase with the visibility of the event-locked scalp response (Fig. 3B). Given the small number of subjects, this relationship was interpreted descriptively.

### 3.3. Intracranial Waveform Reconstruction: “What”

In contrast to the subject-dependent where results, waveform reconstruction was strongly constrained by the patient-specific template (Fig. 3C, Table 1). Across subjects, the unweighted mean waveform correlation was 0.752 for the model and 0.754 for the fixed template, corresponding to a mean model-template difference of only −0.002. At the subject level, differences ranged from −0.019 in sub-02 to +0.019 in sub-08, and all 95% confidence intervals for the model-template difference included zero. Thus, the scalp-conditioned model did not consistently improve full-waveform reconstruction beyond the fixed patient-specific template.

Residual-waveform prediction showed similarly heterogeneous and generally modest performance. Mean residual correlation ranged from −0.020 to 0.378; confidence intervals excluded zero only in sub-06 and sub-08. Across subjects, model waveform correlation tended to be higher when the annotated IEDs were more stereotyped, as quantified by the median leave-one-out waveform similarity (Fig. 3D). The proportion of opposite-polarity events is additionally shown in the same panel. This exploratory relationship, together with the small model-template differences, indicates that the apparent success of waveform reconstruction was largely determined by how reproducible the characteristic intracranial waveform was within each patient.

## 4. Discussion

This study examined two distinct aspects of intracranial IED information potentially recoverable from scalp EEG: where the event occurs in time within a local IED-containing segment and what intracranial waveform morphology is present at that event. The analyses yielded different results for these questions. Temporal localization was clearly above chance in several subjects and was degraded by input ablation or temporal scrambling, supporting a genuine contribution from scalp EEG. In contrast, waveform reconstruction was generally similar to the fixed patient-specific template, and prediction of event-specific residual morphology was modest. These findings indicate that scalp-recoverable information is not uniform across tasks or patients: temporal information can be recoverable when the scalp manifestation is sufficiently observable, whereas single-contact intracranial morphology in this dataset is strongly constrained by the patient-specific waveform prior.

The marked intersubject variability provides insight into these differences. Where accuracy tended to be higher in subjects with a more visible event-locked scalp response, consistent with the expected dependence of noninvasive observability on signal strength. In contrast, what performance closely followed the reproducibility of the intracranial template. Thus, a high waveform correlation does not necessarily imply successful recovery of event-specific intracranial information; it may largely reflect that the target waveform itself is highly stereotyped. This distinction is especially important in patient-specific reconstruction problems, in which a strong within-patient prior can produce apparently accurate outputs even when the noninvasive input contributes little. For the where task, input-control analyses provided direct evidence of scalp-dependent temporal information. For the what task, the near-zero improvement over the fixed template and the modest residual correlations instead indicated that most apparent waveform reconstruction was explained by the patient-specific prior.

The present task is complementary to, rather than a replacement for, IED detection. Automated detectors such as SpikeNet2 (Li et al., 2025) address whether and when an IED is present in continuous scalp EEG, which is a prerequisite for practical deployment. Here, the where task begins from a local segment already known to contain the annotated intracranial event and therefore should not be interpreted as continuous IED detection. The what task addresses a different potential clinical use. A patient-specific model could be trained during simultaneous scalp EEG-iEEG monitoring, when the characteristic intracranial IED is directly observed, and subsequently applied to routine noninvasive scalp EEG after electrode explantation. In such longitudinal follow-up, detection could quantify whether and how often IEDs occur, whereas morphology reconstruction could potentially indicate whether the previously characterized intracranial discharge remains stable or changes over time. Such changes may include a shift between a spike-like and a broader sharp-wave morphology and could provide information complementary to event counts alone. The present results, however, show that this clinical application remains prospective, because event-specific morphology was only weakly recovered beyond the fixed patient-specific template.

A further limitation is the asymmetric spatial information in the available dataset. Scalp EEG provides a multichannel spatial field, whereas the reconstruction target is a single representative iEEG contact selected in the original public dataset. Similar waveforms at one intracranial contact do not necessarily imply identical spatial generators across events. Changes in source extent, orientation, or cancellation may alter the scalp field while producing relatively similar activity at the selected contact, thereby limiting one-to-one mapping from multichannel scalp EEG to a single intracranial waveform. Future studies should therefore extend the problem to simultaneous multicontact iEEG and multichannel scalp EEG, ideally incorporating anatomically informed forward models to evaluate both temporal and spatial relationships between invasive and noninvasive activity.

The study has several additional limitations. Only six subjects were available, each model was trained and tested within the same patient, and the dataset contained one characteristic IED type per subject. The released recordings consist of event-centered 2-s epochs rather than continuous EEG, and additional non-annotated spike-like transients within some epochs cannot be reliably classified without the original continuous clinical context. Consequently, non-IED segments, continuous detection performance, cross-patient generalization, and robustness to longitudinal changes in recording conditions could not be evaluated. The exploratory subject-level associations should also be interpreted cautiously because of the small sample size. Larger datasets containing continuous scalp EEG, multiple intracranial contacts, and repeated recordings across time will be required to determine the clinical utility and generalizability of this framework.

Patient-specific scalp EEG contained recoverable information about the temporal location of intracranial IEDs in some subjects, particularly when the event had a clear scalp manifestation. In contrast, reconstruction of single-contact intracranial morphology was largely explained by a fixed patient-specific template, with limited additional event-specific information recovered from scalp EEG. Separating where from what, together with direct input controls, provides a more stringent framework for evaluating future noninvasive-to-intracranial reconstruction models.

## CRediT Authorship Contribution Statement

Teppei Matsubara: Conceptualization, Formal analysis, Investigation, Methodology,

Writing – original draft, Writing – review & editing.

Ryu Koda: Methodology, Software, Writing – review & editing.

Mark Richardson: Supervision, Writing – review & editing.

Steven Stufflebeam: Supervision, Writing – review & editing.

## Funding

This research did not receive any specific grant from funding agencies in the public, commercial, or not-for-profit sectors.

## Ethics Statement

The original study was approved by the local Ethics Committee of Niguarda Hospital, Milan, Italy (protocol 463-092018), and all participants provided informed consent (Zauli et al., 2024). The present study analyzed de-identified data available through the public repository.

## Declaration of Competing Interest

None to declare.

## Data Availability Statement

The dataset analyzed in this study is publicly available in the Open Science Framework repository at https://osf.io/89ndr/ (Zauli et al., 2024).

## Notes

### Competing Interest Statement

The authors have declared no competing interest.

## References

Hu M, Chen J, Jiang S, Ji W, Mei S, Chen L, Wang X. E2SGAN: EEG-to-SEEG translation with generative adversarial networks. Front Neurosci, 2022; 16: 971829.

Janmohamed M, Nhu D, Kuhlmann L, Gilligan A, Tan CW, Perucca P, O’Brien TJ, Kwan P. Moving the field forward: detection of epileptiform abnormalities on scalp electroencephalography using deep learning-clinical application perspectives. Brain Commun, 2022; 4: fcac218.

Jing J, Sun H, Kim JA, Herlopian A, Karakis I, Ng M, Halford JJ, Maus D, Chan F, Dolatshahi M, Muniz C, Chu C, Sacca V, Pathmanathan J, Ge W, Dauwels J, Lam A, Cole AJ, Cash SS, Westover MB. Development of Expert-Level Automated Detection of Epileptiform Discharges During Electroencephalogram Interpretation. JAMA Neurol, 2020; 77: 103–8.

Kural MA, Duez L, Sejer Hansen V, Larsson PG, Rampp S, Schulz R, Tankisi H, Wennberg R, Bibby BM, Scherg M, Beniczky S. Criteria for defining interictal epileptiform discharges in EEG: A clinical validation study. Neurology, 2020; 94: e2139–e47.

Li J, Goldenholz DM, Alkofer M, Sun C, Nascimento FA, Halford JJ, Dean BC, Galanti M, Struck AF, Greenblatt AS, Lam AD, Herlopian A, Nwankwo C, Weber D, Maus D, Haider HA, Karakis I, Yoo JY, Ng MC, Selioutski O, Taraschenko O, Osman G, Katyal R, Schmitt SE, Benbadis S, Cash SS, Tatum WO, Sheikh Z, Kong WY, Bayas G, Turley N, Hong S, Westover MB, Jing J. Expert-Level Detection of Epilepsy Markers in EEG on Short and Long Timescales. NEJM AI, 2025; 2.

Pyrzowski J, Le Douget JE, Fouad A, Sieminski M, Jedrzejczak J, Le Van Quyen M.Zero-crossing patterns reveal subtle epileptiform discharges in the scalp EEG. Sci Rep, 2021; 11: 4128.

Ramantani G, Maillard L, Koessler L. Correlation of invasive EEG and scalp EEG. Seizure, 2016; 41: 196–200.

Rosenow F, Luders H. Presurgical evaluation of epilepsy. Brain, 2001; 124: 1683–700.

Spyrou L, Martin-Lopez D, Valentin A, Alarcon G, Sanei S. Detection of Intracranial Signatures of Interictal Epileptiform Discharges from Concurrent Scalp EEG. Int J Neural Syst, 2016; 26: 1650016.

Tao JX, Ray A, Hawes-Ebersole S, Ebersole JS. Intracranial EEG substrates of scalp EEG interictal spikes. Epilepsia, 2005; 46: 669–76.

Vaswani A, Shazeer N, Parmar N, Uszkoreit J, Jones L, Gomez AN, Kaiser Ł, Polosukhin I. Attention is all you need. Advances in neural information processing systems, 2017; 30.

Yamaguchi T, Uehara T, Okadome T, Mukaino T, Shimogawa T, Mukae N, Shigeto H, Isobe N. Scalp Electroencephalography Markers of Hidden Interictal Epileptiform Discharges in the Mesial Temporal Lobe. Int J Neural Syst, 2026; 36: 2650002.

Zauli FM, Del Vecchio M, Pigorini A, Russo S, Massimini M, Sartori I, Cardinale F, d’Orio P, Mikulan E. Localizing hidden Interictal Epileptiform Discharges with simultaneous intracerebral and scalp high-density EEG recordings. J Neurosci Methods, 2024; 409: 110193.

